# NGLY1 Deficiency: A Prospective Natural History Study (NHS)

**DOI:** 10.1101/2023.05.10.23289581

**Authors:** Sandra Tong, Pamela Ventola, Christina H. Frater, Jenna Klotz, Jennifer M. Phillips, Srikanth Muppidi, Selina S. Dwight, William F. Mueller, Brendan J. Beahm, Matt Wilsey, Kevin J. Lee

## Abstract

N-glycanase 1 (NGLY1) Deficiency is a debilitating, ultra-rare autosomal recessive disorder caused by loss of function of NGLY1, a cytosolic enzyme that deglycosylates other proteins. It is characterized by severe global developmental delay and/or intellectual disability, hyperkinetic movement disorder, transient elevation of transaminases, (hypo)alacrima, and progressive, diffuse, length-dependent sensorimotor polyneuropathy. A prospective natural history study (NHS) was conducted to elucidate clinical features and disease course. Twenty-nine participants were enrolled (15 onsite, 14 remotely) and followed for up to 32 months, representing ∼29% of the ∼100 patients identified worldwide. Participants exhibited profound developmental delays, with almost all developmental quotients below 20 on the Mullen Scales of Early Learning, well below the normative score of 100. Increased difficulties with sitting and standing suggested decline in motor function over time. Most patients presented with (hypo)alacrima and reduced sweat response. Pediatric quality of life was poor except for emotional function. Language/communication and motor skill problems including hand use were reported by caregivers as the most bothersome symptoms. Levels of the substrate biomarker, GlcNAc-Asn (aspartylglucosamine; GNA), were consistently elevated in all participants over time, independent of age. Liver enzymes were elevated for some participants but improved especially in younger patients and did not reach levels indicating severe liver disease. Three participants died during the study period. Data from this NHS informs selection of endpoints and assessments for future clinical trials for NGLY1 Deficiency interventions. Potential endpoints include GNA biomarker levels, neurocognitive assessments, autonomic and motor function (particularly hand use), (hypo)alacrima, and quality of life.

## Introduction

*N*-glycanase 1 (NGLY1) Deficiency (OMIM <u>615273</u>) is a severely debilitating, ultra-rare, autosomal recessive neurodevelopmental disorder characterized by five core clinical features: global developmental delay and/or intellectual disability, a hyperkinetic movement disorder, transient elevation of transaminases, (hypo)alacrima, and a progressive, diffuse, length-dependent sensorimotor polyneuropathy (Adams 2018; Enns 2014; Lam 2018). In addition to these five core features, individuals may experience seizures, reduced ability to sweat, skeletal problems such as contractures, scoliosis, and skeletal deformities, hearing loss, vision impairment, and adrenal insufficiency (Lam 2018). There are approximately 100 individuals living with NGLY1 Deficiency identified globally (Pandey 2022). Because so few cases have been identified, estimated lifespan is unknown. While some individuals survive into early adulthood, there have been reports of death during infancy and adolescence (Lam 2018, Dabaj 2021, Kalfon 2021, Stuut 2021). Most affected individuals require constant daily care due to difficulty performing activities such as feeding, toileting, and bathing, and many are unable to walk or speak (Lam 2018).

NGLY1 Deficiency is caused by mutations in the *NGLY1* gene that encodes the cytosolic protein *N*-glycanase 1, or NGLY1 (Enns 2014). NGLY1 cleaves *N*-glycans from proteins targeted for degradation as part of the endoplasmic reticulum-associated degradation (ERAD) pathway. ERAD is a cellular quality control pathway in which misfolded or improperly processed proteins in the endoplasmic reticulum (ER) are translocated to the cytosol and delivered to the proteasome for degradation. NGLY1 cleaves N-linked glycans from ERAD substrate glycoproteins by hydrolyzing the proximal N-acetylglucosamine-asparagine (GlcNAc-Asn) amide bond prior to degradation by the proteasome. This cleavage results in two products, a free oligosaccharide and a deaminated protein (Suzuki 2016). In the absence of NGLY1, glycans can be cleaved from the protein at alternate positions within the glycan, resulting in an incompletely deglycosylated protein that retains the proximal GlcNAc bound to Asn. This protein can then be degraded to peptides, which are hydrolyzed to amino acids. Because the substrate GlcNAc-Asn bond normally cleaved by NGLY1 remains intact, GlcNAc-Asn (GNA) accumulates in the cytosol in cells that lack NGLY1 activity (Haijes 2019; Mueller 2022). As a consequence, GNA levels are consistently elevated in NGLY1 deficient organisms and cells (Mueller 2022) and can be measured using a quantitative assay in dried blood spots, urine, and plasma samples from NGLY1 Deficiency patients (Haijes 2019; Mueller 2022).

Restoration of NGLY1 activity in NGLY1 deficient cellular and animal models leads to a reduction of GNA levels. Thus measurement of GNA concentration directly reflects NGLY1 activity and could be expected to predict clinically meaningful restoration of NGLY1 activity in a therapeutic setting (Haijes 2019; Mueller 2022; Zhu 2022). A reliable biomarker for NGLY1 Deficiency is critical given the small population of affected individuals, the slowly progressive nature of the disease, and the high degree of phenotypic variability (Mueller 2022).

The specific factors underlying the heterogenous presentation of NGLY1 Deficiency are not known. Diagnosis of NGLY1 Deficiency is based on a combination of genetic testing, a detailed medical history, and clinical examination to identify key clinical features, particularly (hypo)alacrima, which is only present in very few disorders. Increased levels of plasma or urine GNA may support a diagnosis of NGLY1 Deficiency when genetic testing shows variants of uncertain significance (Haijes 2019; Mueller 2022). Given the rarity of the disease and its heterogenous nature, there is often a significant delay between onset of symptoms and diagnosis (Enns 2014, Adams 2018).

There are no approved treatments for NGLY1 Deficiency; rather, symptoms must be managed daily by caretakers. Symptomatic treatments include feeding tubes to provide nutrients; lubricating eye drops or ointments for dry eyes related to poor tear production; anti-seizure medication; occupational, physical, and speech therapy to assist with activities of daily living; and psychosocial support for families and caregivers (Lam 2018; Lam 2017). Nearly half of patients also require invasive procedures or surgery for musculoskeletal issues associated with NGLY1 Deficiency (Cahan 2019).

To support the development of disease-modifying therapies, a full elucidation of the natural history and clinical manifestations of NGLY1 Deficiency is needed. As the disorder was recently identified (Need 2012), a prospective natural history study (NHS) that enrolled a substantial portion of the known affected individuals was conducted to improve understanding of the presentation and course of the disorder.

This paper reports on the findings of the NGLY1 Deficiency prospective NHS described above (NCT03834987). The objectives of this study were to understand the clinical spectrum and progression of NGLY1 Deficiency using standardized clinical and neurodevelopmental assessments, and to identify and define clinical and biomarker endpoints for use in therapeutic trials.

## Materials and Methods

### Study design

The NGLY1 NHS reported here was a prospective, longitudinal observational study conducted by Stanford University (http://clinicaltrials.gov, NCT03834987) and funded by the Grace Science Foundation (gracescience.org), a research and advocacy group for NGLY1 Deficiency. The planned enrollment was up to 50 patients with a confirmed diagnosis of NGLY1 Deficiency, with up to 15 time points per participant. Study procedures included annual in-person (“onsite” participants) or annual remote (“remote” participants) evaluations along with remote assessments for each group every 4-months between annual evaluations. Established developmental and health-related quality of life (HRQoL) measures as well as standardized laboratory and clinical assessments were employed.

### Participants

Participants included males and females of any age with a suspected or confirmed diagnosis of NGLY1 Deficiency based on the identification of (likely) pathogenic variants in both *NGLY1* alleles and clinical characteristics consistent with the disease. Parent(s)/legal representative(s) of participants were required to give informed consent/assent for study participation; for the results of the study to be presented at scientific/medical meetings or published in scientific journals; and to be willing for the participant to provide clinical data, provide biological samples, and participate in standardized assessments. Willingness to travel to Palo Alto, CA was favored, but not required. Each study participant’s data records were assigned a unique code number. Information about the code was kept in a secure location and access was limited to key research study personnel.

Participants were excluded from the study if they had a second, confirmed disorder affecting neurodevelopment or with overlapping symptoms of NGLY1 Deficiency.

Participants were permitted to be enrolled as “onsite” or “remote” participants. Onsite participants were scheduled to visit the clinical site once per year for the study duration, with 2 telephone / video visits in between each annual visit at 4-month intervals. Remote participants were scheduled for annual telephone / video visit with 2 telephone / video visits in between each annual visit at 4-month intervals. Most assessments began at baseline, except for SSDS, Schirmer’s test, and the question about the most bothersome symptom, which began in Year 2. These assessments therefore have a lower number of data points.

Participants enrolled in the study at different times (between February 15, 2019 and October 14, 2020); however, the end date of the study was the same for all patients. For this reason, the number of participants with an assessment at each study time point was variable and generally decreased over time. This effect was compounded by the COVID-19 pandemic, which caused delays in enrollment and assessments.

### Assessments

#### Primary Objectives

There were two primary outcome measures for the NGLY1 Deficiency NHS: 1) developmental assessment at baseline and longitudinally, as measured by the established scales, and 2) disease course over time as measured by standardized medical histories.

The following established scales were used for the primary outcome capturing development assessment: Mullen Scales of Early Learning (MSEL), Vineland Adaptive Behavior Scales, 3^rd^ Edition (Vineland-3), Peabody Scales of Motor Development (PDMS-2), and Beery-Buktenica Test of Visual Motor Integration (Beery VMI).

The MSEL is a direct assessment and provides a broad global assessment of development in children from birth to 68 months by examining 5 key subscales: gross motor, fine motor, visual reception, expressive language, and receptive language. The developmental quotient (DQ), which is calculated by averaging the 5 areas of the Mullen scale and dividing the developmental age by the chronological age x 100 (a DQ of 100 corresponds to an exact match between the developmental and chronological age), provides an overall approximation of the affected individual’s ability to function (Aylward 2008).

The Vineland-3 is a parent/caregiver-reported measure that uses a semi-structured interview to assess adaptive behavior across four domains: Communication, Daily Living Skills, Socialization, and Motor Skills. It is used to support the diagnosis of intellectual and developmental disabilities (Farmer 2020). The Vineland Adaptive Behavior Composite is derived from the Communication, Daily Living Skills, and Socialization domains.

The PDMS-2 includes five subscales used to calculate 2 quotients: Grasping and Visual–Motor Integration (which are used to calculate the Fine Motor Quotient [FMQ]); and Stationary, Locomotion and Object Manipulation, replaced by the Reflexes subtest for children up to eleven months old (which are used to calculate the Gross Motor Quotient [GMQ)]). FMQ and GMQ can also be combined to yield a Total Motor Quotient (TMQ). FMQ, GMQ, and TMQ are scored similarly to the DQ. They compare developmental age to chronological age; a score of 100 represents an exact match between developmental age and chronological age. Scores that are lower or higher than 100 represent a developmental age that is lower or higher than the chronological age, respectively (Rebelo 2021).

The Beery VMI is administered by a clinician and assesses the ability to integrate visual and motor activities in individuals 2 years of age and older. It requires individuals to copy a sequence of geometric forms using paper and pencil. While the Beery VMI was a planned assessment for this NHS, participants in the study were too developmentally impaired to attempt this test (Spencer 2010).

The Bruininks-Oseretsky Test of Motor Proficiency Second Edition (BOT-2) and Differential Ability Scales-II (DAS-2) were used for higher functioning subjects but are not discussed in this paper because they were only conducted in 3 participants.

The primary outcome measure of disease course over time was measured by collecting standardized medical histories that included general physical evaluations, clinical neurologic evaluations, standardized dysmorphology evaluations, laboratory studies (comprehensive metabolic panel, liver function tests, creatine kinase, lactic acid, fasting lipid panel, adrenocorticotropic hormone [ACTH], cortisol, biochemical studies), scoring of movement disorders (including 10-meter walk test [10-MWT]), and standardized and Schirmer’s ophthalmologic evaluations.

Schirmer’s test was used to evaluate (hypo)alacrima. This test determines whether the eye produces sufficient tears by placing calibrated test strips of non-toxic filter paper within the lower eyelid of each eye for 5 minutes and measuring the distance traveled by the tears on the test strips (Karampatakis 2010).

The sitting and standing abilities were part of the neurologic exam and used to measure gross motor function (Pereira 2016). Minimal support was defined as hands only, and maximal support was the requirement for truncal support. The 10-MWT was used to measure walking speed in meters per second over a short walking distance at an individual’s usual walking speed (comfortable speed) and/or walking as quickly as possible (fast speed) (de Baptista 2020).

#### Secondary Objectives

Secondary objectives included identification of clinical endpoints or biomarkers for therapeutic trials, and caregiver and participant QoL at baseline and longitudinally. Participant QoL was measured through the Pediatric Quality of Life Inventory (PedsQL) as reported by the caregiver. The PedsQL measures quality of life in children and adolescents; it includes 23 questions in 4 domains: physical, emotional, social, and school functioning. Caregiver QoL was measured through self-report using SF-36. The SF-36 measures quality of life in adults, adolescents, and children and includes 36 questions in 8 domains: vitality, physical functioning, bodily pain, general health perceptions, physical role functioning, emotional role functioning, social role functioning, and mental health. Finally, a single question asking caregivers to identify the most bothersome symptom for the participant was included.

Other procedures completed at baseline and longitudinally as tolerated included: electroencephalogram (EEG), seizure diary (when applicable for those experiencing active seizures), nerve conduction studies, and quantitative studies of autonomic function (QSART). EEG and seizure diary results are reported in a separate paper (Levy 2022).

#### Biomarker

The assay for GNA substrate biomarker has been described previously (Mueller 2022). This assay uses liquid chromatography/tandem mass spectrometry (LC/MS/MS) in positive electrospray ionization mode (ESI+) for the quantitation of GNA in human plasma. Test samples, calibration standards, and QC samples were processed by protein precipitation with acetonitrile to enrich the analyte in the matrix samples. Samples were homogenized in phosphate-buffered saline (PBS) and mixed with three volumes of ice cold Internal Standard Solution (acetonitrile containing 60 ng/ml d3-GNA, Omicron Biochemicals, Inc.; catalog number AAG-004, Lot# Q01-N0816). They were then centrifuged at 6100 g for 30 min. An aliquot of each supernatant was transferred to an autosampler plate. The supernatant was separated via HPLC (Shimadzu VP Series 10 System) and analyzed via MS/MS (Applied Biosystems/MDS SciEx API 4000). Detection and accuracy were assessed in surrogate matrices (PBS + bovine serum albumin [BSA], charcoal stripped serum). Each surrogate matrix was spiked with 30 and 300 ng/ml of GNA (Omicron Biochemicals, Inc.; catalog number AAG-003), processed and analyzed to determine recovery and accuracy. The processed samples were analyzed by LC/MS/MS using a HILIC column. GNA was quantified using d3-GNA (β-D-GlcN[2H3]Ac-(1,N)-Asn) as an internal standard (IS). The calibration range was 5 to 2500 ng/mL using 20 μL of sample. The analyte/IS peak area ratios (y) versus the nominal analyte concentrations (x) of the calibration samples were used to fit a calibration curve by power regression (origin excluded). The analyte concentrations for the calibration standards, quality control samples, and unknown (study) samples were calculated using the established calibration equation.

### Statistical methodology

This NHS was descriptive; thus, data were considered hypothesis-generating and not subjected to power analysis. Demographic data and baseline disease characteristics were summarized for all enrolled participants. Clinical and biological data were summarized over the study period. Data were presented descriptively, and regression and/or ANOVA analyzes were performed to characterize disease course and any associated markers.

GNA biomarker data was analyzed using R (version 3.5.1 (2018-07-02) -- “Feather Spray”) Scripts available upon request.

## Results

### Participant Disposition and Demographics

The study enrolled 29 participants with a confirmed diagnosis of NGLY1 Deficiency,15 onsite participants and 14 remote participants. The data presented here expand on the baseline characteristics and on the disposition of those reported by Levy 2022. Three participants died during the study period (Table 1). All other patients remained in the study throughout its duration. Participants were followed for a median of 23 months, with a range of 8-32 months. Because participants entered the study at different times, they were followed for varying lengths of time up to the study end date. Thus, the number of patients at each time point is provided when describing the results.

**Table 1:** Participant Disposition and Demographics

| Demographic |  |
| --- | --- |
| Number of participants – n | 29 |
| - Onsite – n (%) | 15 (51.7) |
| - Remote – n (%) | 14 (49.3) |
| Discontinuations during study – n (%) | None |
| **Demographic** |  |
| Deaths during study – n (%) | 3 (10.3) |
| Age at death – age range (number) | 10-15 years (2); 20-25 years (1) |
| Age at: mean (SD)[range] – years |  |
| - Study entry | 9.93 (6.279)[1-27] |
| - Diagnosis | 7.1 years (5.45)[0 – 16.1] |
| Gender – n (%) | 29 (100%) |
| - Male | 15 (51.7) |
| - Female | 14 (49.3) |
| Weight as Z-score - n (%) | 15 (52%) |
| Mean (SD) [range] | -2.1 (0.97) [-0.57 to -3.97] |

The mean age [range] of participants was 9.93 [1-27] years at entry and 7.1 [0-16.1] years at diagnosis. The mean weight [range] expressed as Z-score was −2.08 [−0.57 to −3.97], thus indicating for all participants body weights below the norm for age.

### Developmental Assessments

#### Mullen Scales of Early Learning (MSEL)

Participants who were given the MSEL had profound global developmental delays, with most developmental quotient (DQ) scores below 20, indicating a developmental age much lower than the normative score of 100 +/− 15 (**Figure *1***). No participant showed improvement in DQ over time. Generally, the DQ scores were slightly declining over time for individual participants.

**Figure 1:**
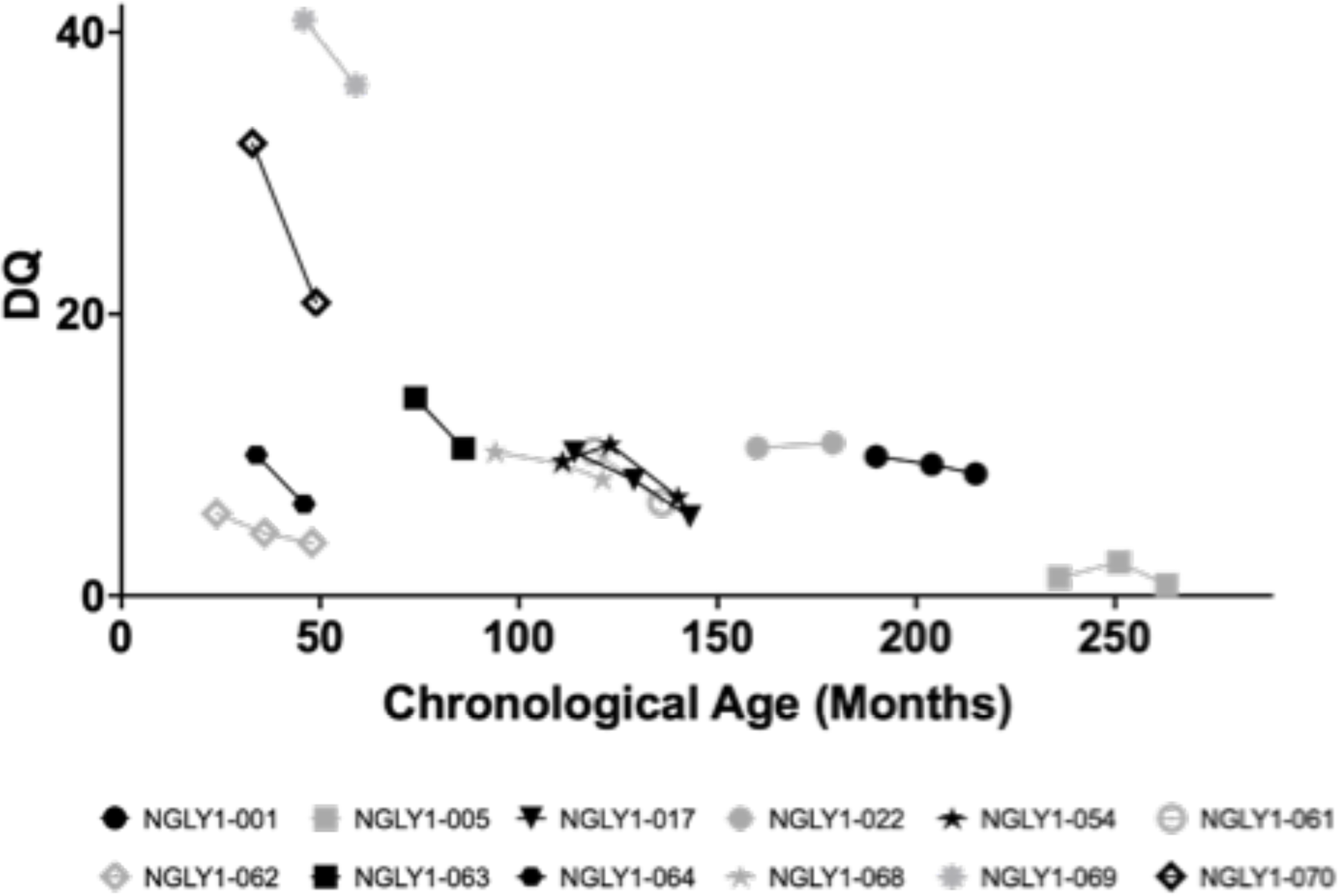
Scatter Plot of Mullen-DQ vs Chronological Age. Legend: Individual participant DQ scores over time are shown. The normative value is 100 +/− 15. Black symbols are female and gray symbols are male subjects.

All MSEL subscales were also profoundly impaired and generally without remarkable improvement; age equivalents were lowest in expressive language and highest in visual reception (data on file).

Almost all participants had profound global impairment in adaptive functioning at baseline as reflected in the Vineland Adaptive Behavior Composite (ABC) standard score where a value of 100 (SD 15) is considered normal (Figure 2). The baseline mean (SD) was 49.43 (22.076) with a range of 23.0-88.0 (n=14). Annual year 2 showed mean (SD) of 40.80 (16.825) with a range of 23.0-71.0 (n=10), which was indicative of no improvement. Participants showed global impairment in all domains of the Vineland Adaptive Behavior Scales (data on file). While there were some fluctuations over time, the magnitudes of change were neither statistically significant nor clinically meaningful; participants generally remained severely impaired. Two participants, subjects were higher functioning in several subdomains relative to other participants.

**Figure 2:**
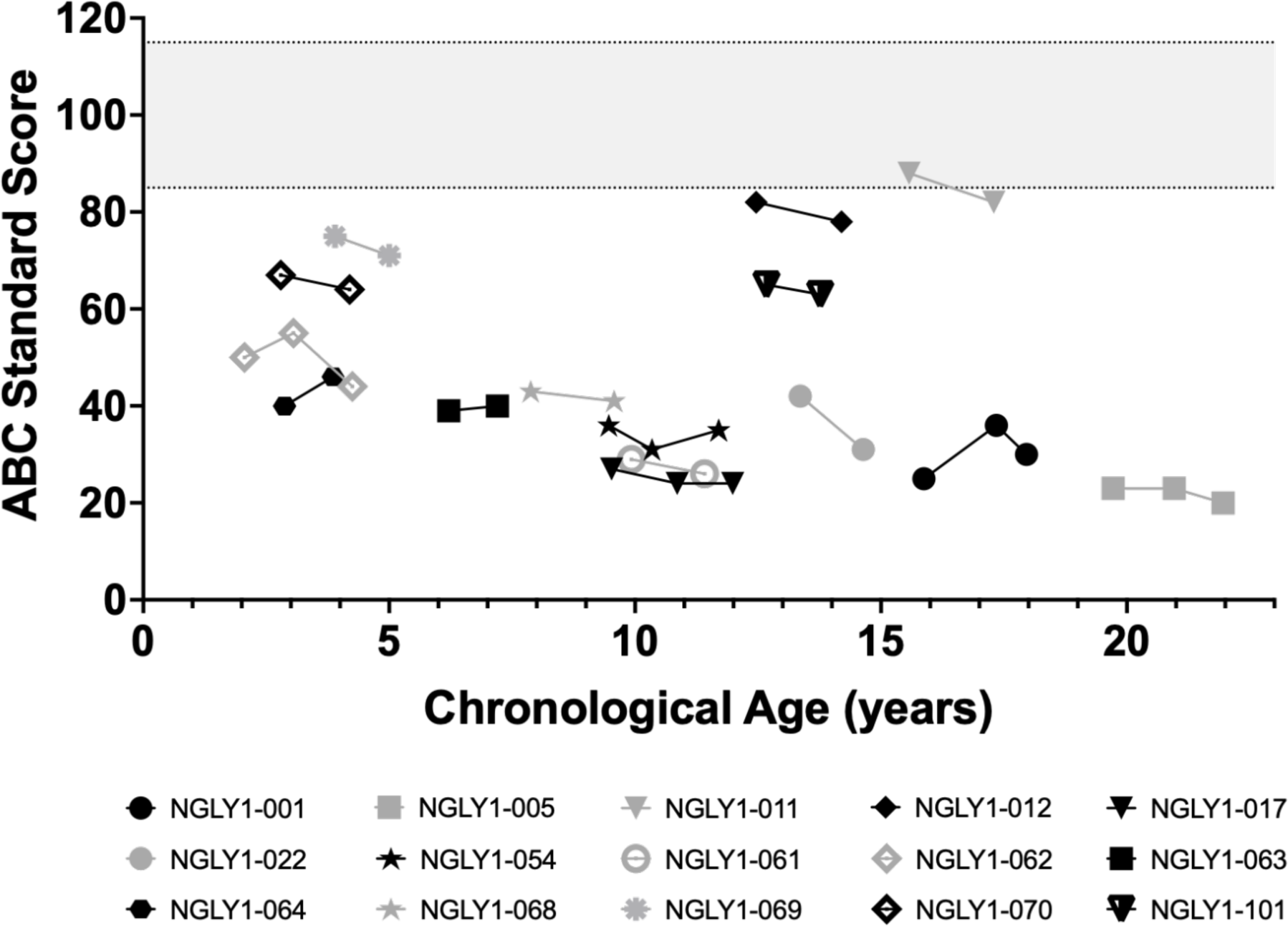
Vineland ABC. Legend: Individual participant Vineland ABC scores over time are shown. The normative value is 100 +/− 15 (shaded area). Black symbols are female and gray symbols are male subjects.

#### Peabody Developmental Motor Scales – 2^nd^ Edition (PDMS-2)

Participants assessed for the Peabody Developmental Motor Scales – 2^nd^ Edition (PDMS-2) had clinically significant impairments in all subscales of gross and fine motor function, The PDMS-2 provides a more detailed analysis of gross and fine motor skills in children from birth to 71 months of age. All participants who could be assessed with PDMS-2 showed marked impairment in DQ for Gross Motor (Figure 3A) and Fine Motor (Figure 3B) function and had clinically significant impairments in all subscales of gross and fine motor function. For example, Grasping age equivalence was at or below 8 months and Object Manipulation age equivalence was below or at 20 months (data on file), even though chronologic age was as high as 260 months (∼21 years old). These data illustrate a profound impairment in upper extremity function in study participants. Profound motor impairment was also observed in all other PDMS-2 domains (data on file). While there were small changes over time for many participants, the profound motor impairments were generally stable or worsening over time.

**Figure 3:**
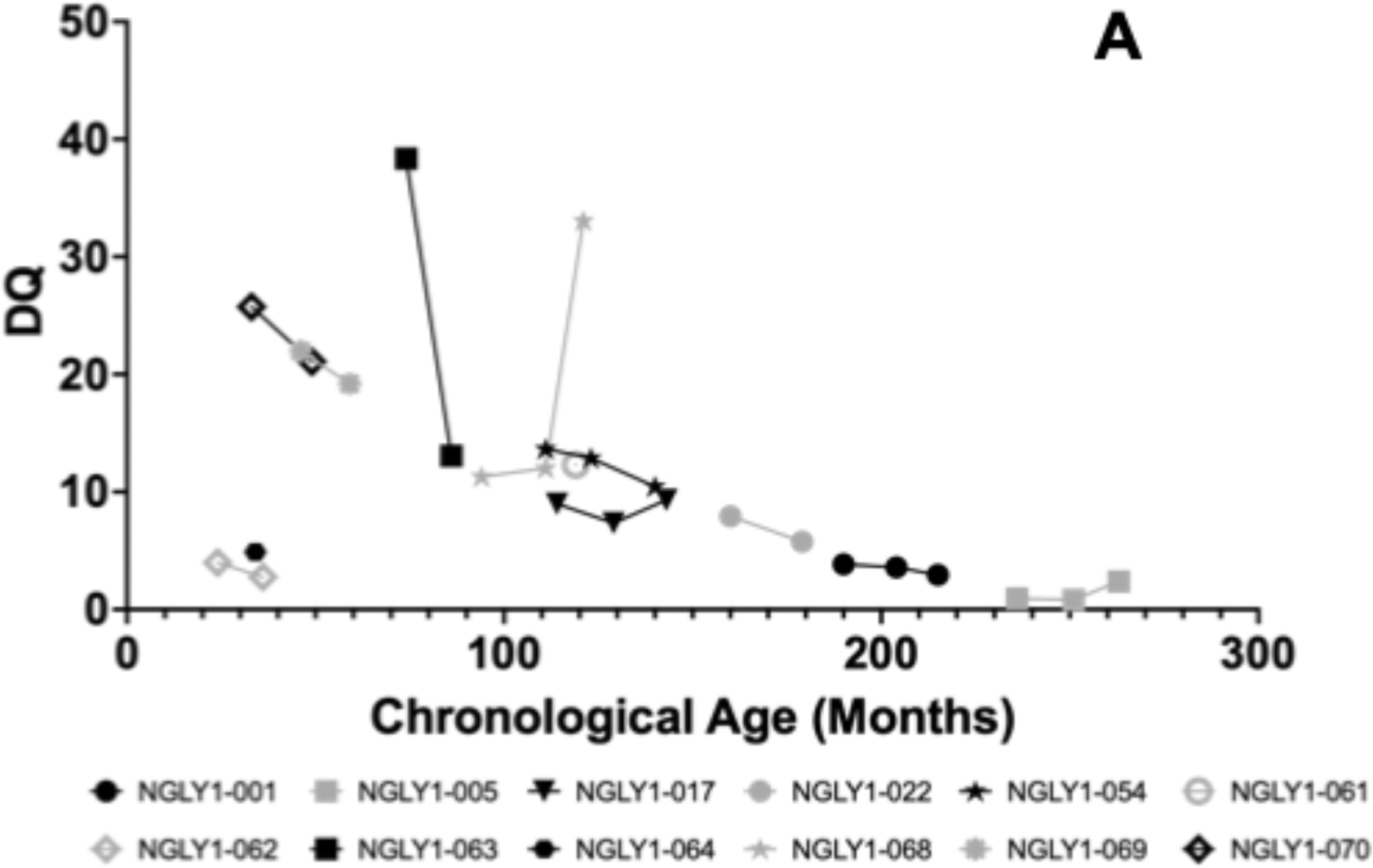
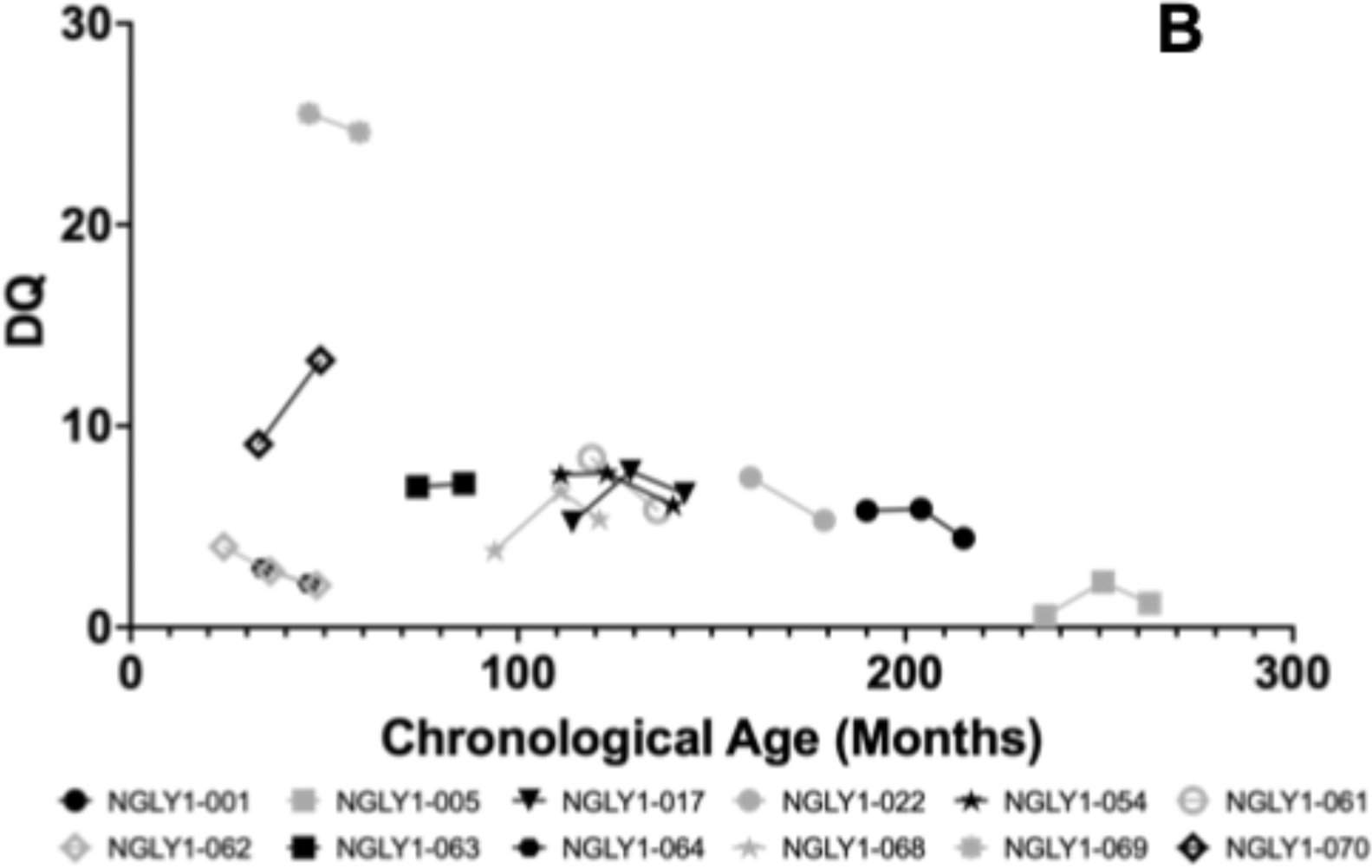
PDMS-2 Gross Motor (A) and Fine Motor (B) Function. Legend: Individual participant PDMS-2 Gross Motor (A) and Fine Motor (B) Function scores over time are shown. The normative values are 100 +/−15. Black symbols are female and gray symbols are male subjects.

### Clinical Disease Course Over Time

#### Motor Function Parameters (Sitting and Standing Ability)

Motor function was significantly impaired and showed progression over time (Table 2). At baseline, 92.3% of participants were able to sit unaided, but by Year 2 this percentage declined to 60.0%. Correspondingly, the proportion of participants requiring support or who were unable to sit increased by Year 2. Less than half of participants (38.5%) were able to stand unaided at baseline, and the percentage declined further at Year 2 to 26.7%. Additionally, the proportion of participants requiring maximal support or unable to stand increased by Year 2. Review of individual participant data confirmed that the worsening trends were due to several participants whose function declined over time (Figure 4).

**Figure 4:**
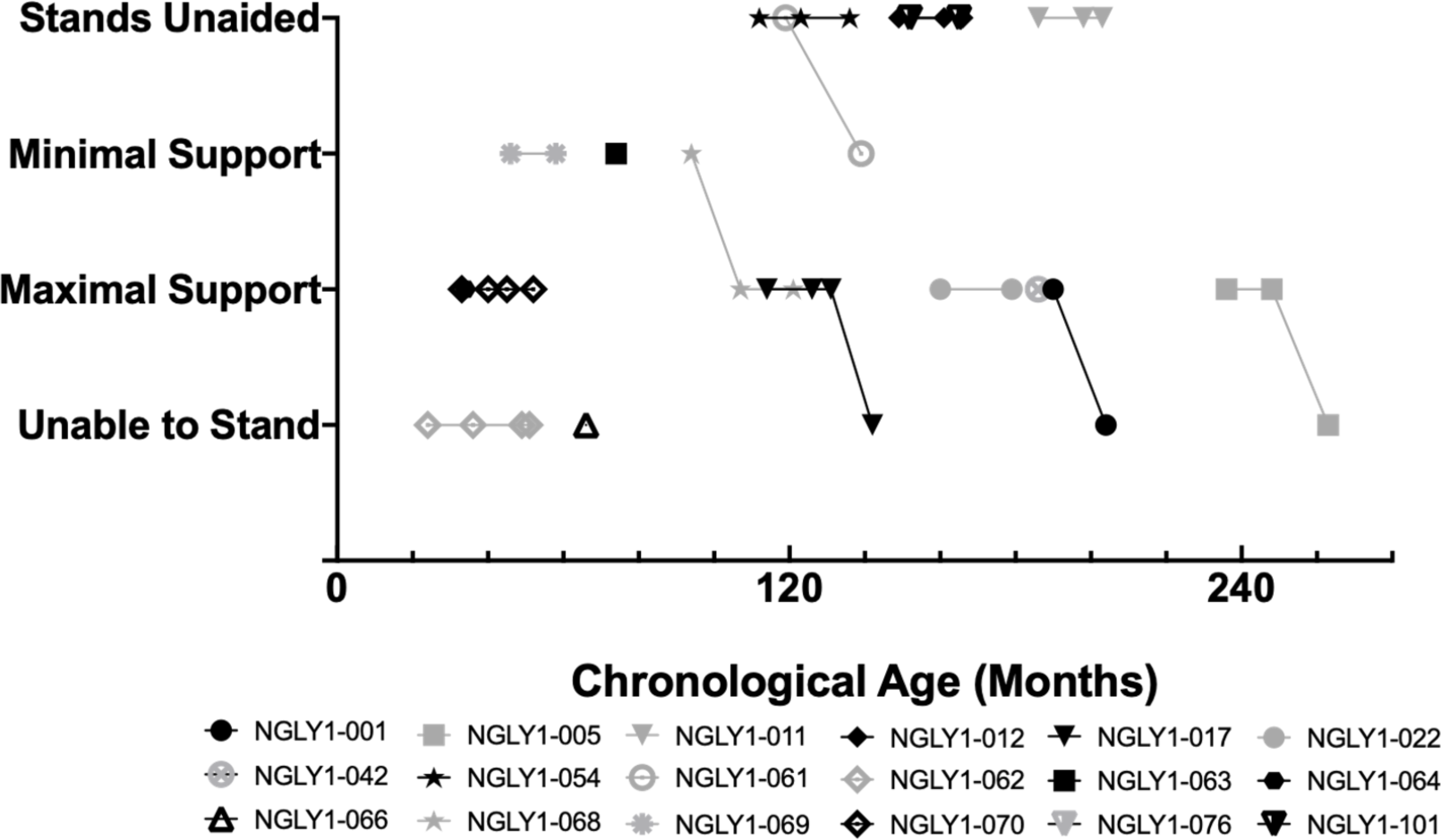
Motor Function: Standing Ability. Legend: Individual participant standing ability over time is shown. Black symbols are female and gray symbols are male subjects.

**Table 2:** Motor Function Parameters

| Parameter | Timeframe* |  |
| --- | --- | --- |
|  | Baseline n/N (%) | First Annual n/N (%) |
| **Sitting Ability** |  |  |
| Sits unaided | 12/13 (92.3) | 9/15 (60.0) |
| Sits with minimal support | 0/13 (0.0) | 4/15 (26.7) |
| Sits with maximal support | 0/13 (0.0) | 1/15 (6.7) |
| Unable to sit | 1/13 (7.7) | 1/15 (6.7) |
| **Standing Ability** |  |  |
| Stands unaided | 5/13 (38.5) | 4/15 (26.7) |
| Stands with minimal support | 2/13 (15.4) | 2/15 (13.3) |
| Stands with maximal support | 5/13 (38.5) | 7/15 (46.7) |
| Unable to stand | 1/13 (7.7) | 2/15 (13.3) |

#### Ophthalmic Evaluations

The Schirmer’s test was used to evaluate (hypo)alacrima using calibrated filter paper to measure tear production. As shown in Figure 5, the mean distance traveled for tears in the Schirmer’s test was generally below normal (≤10 mm; Karampatakis 2010) and for many particpants was near or below the threshold considered strongly positive for (hypo)alacrima (≤ 5 mm) (Li 2012). Because Schirmer’s test was introduced late in the study, many patients had only a single time point collected; longitudinal data for those participants tested more than once showed no consistent change over time. Results were generally similar between left and right eyes (data on file).

**Figure 5:**
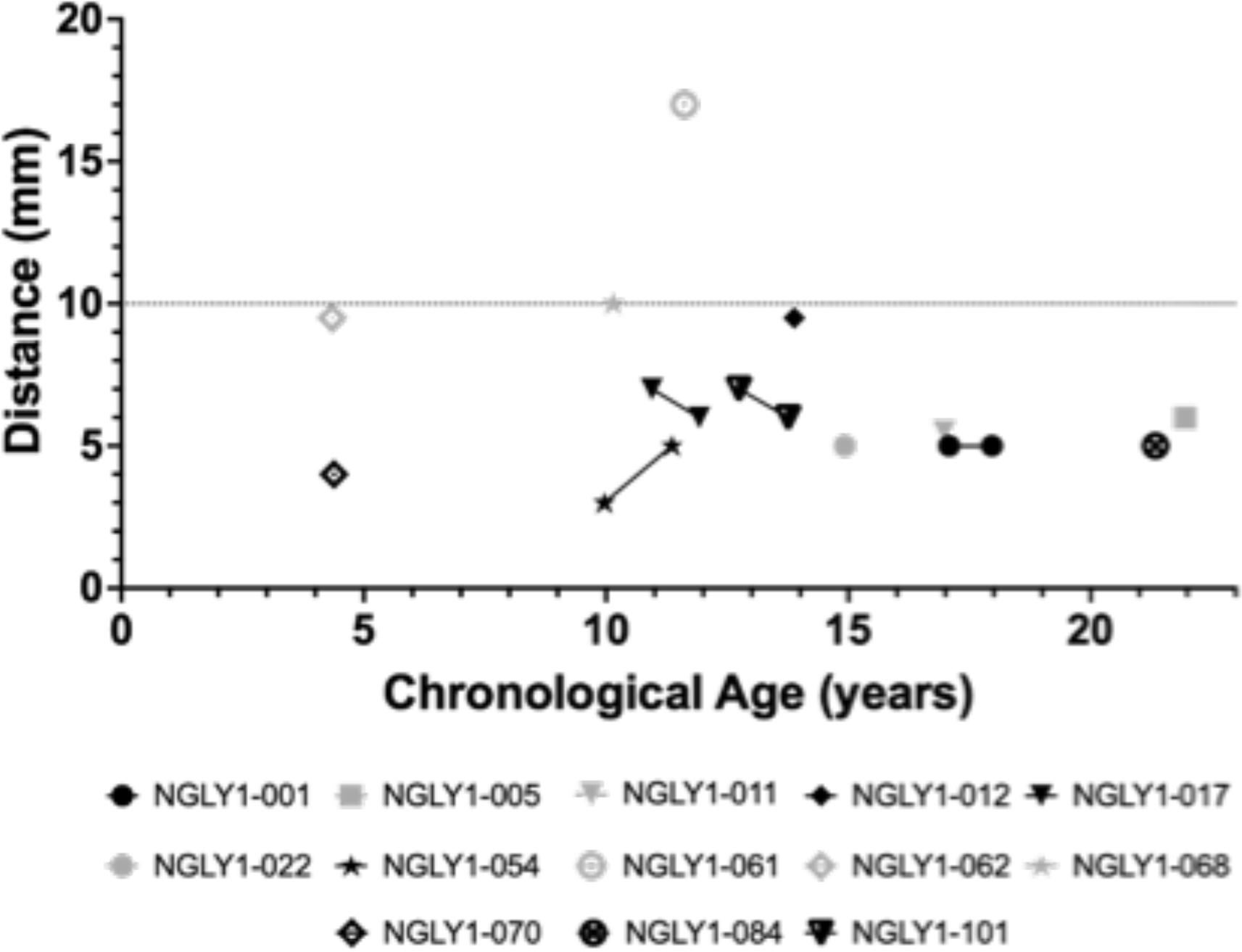
Schirmer’s Lacrimation Test. Legend: Results from left eye. Wetting of ≤ 5 mm per 5 minutes is considered a strong positive test for (hypo)alacrima (Li et al 2012). Greater than 10 mm is considered normal (gray line).

### Other Endpoints

#### 10-Meter Walk Test (10-MWT)

Only half of participants were able to attempt the 10-MWT twice or more at the comfortable speed at baseline, and the majority (84.6%) were not able to attempt the fast speed trial. As there were too few participants for the data to be clinically meaningful, the results are not shown here.

#### Nerve Conduction Studies (NCS)

Interpretation of motor and sensory conduction studies showed a mixed polyneuropathy for most patients. Nerve conduction velocity of the median nerve for motor and sensory nerves generally showed values below normal values reported in the literature (Ryan 2019). Of the 18 participants with NCS assessment, 8 (44%) had at least one median nerve assessment that was not measurable (data on file).

#### Quantitative Measures of Autonomic Function: Quantitative Sudomotor Axon Reflex Test (QSART)

The Quantitative Sudomotor Axon Reflex Test (QSART) measures postganglionic sympathetic sudomotor sweat response. Decreased sweat volume and increased latency (time to onset of sweating) is indicative of peripheral autonomic neuropathy. Participants showed a decreased sweat response (Table 3), with forearm latency and volume shown as a representative location. Mean sweat latency in the forearm was in the high normal to elevated range, indicating delayed sweating (normal values for sweat latency are 60-120 seconds; Illigens 2009). Mean sweat volume in the forearm was in the low normal range at baseline and worsened to below normal at Year 2 (normal values for sweat volume are 0.38-2.86 µL in men and 0.10-1.99 µL in women; Novak, 2011). Other body locations showed generally similar findings (data on file).

**Table 3:** Summary Statistics of QSART

| QSART Domain | Timeframe* |  |
| --- | --- | --- |
|  | Baseline Year 1 | Annual Year 2 |
| **Forearm Latency (sec)** |  |  |
| n | 10 | 5 |
| Mean (SD) | 109.0 (52.05) | 173.0 (81.00) |
| Median | 100.5 | 173.0 |
| Min, Max | 20, 177 | 68, 290 |
| **Forearm Sweat Volume (μL)** |  |  |
| n | 10 | 5 |
| Mean (SD) | 0.5364 (0.45795) | 0.0801 (0.05811) |
| Median | 0.4363 | 0.0490 |
| Min, Max | 0.047, 1.638 | 0.042, 0.179 |

### Quality of Life

Caregiver self-reports for the caregiver’s QoL per SF-36 did not show change over time (data on file).

#### Pediatric Quality of Life Inventory (PedsQL)

The Pediatric Quality of Life Inventory (PedsQL) is a measure of pediatric HRQoL (Hullmann 2011). Total scores for participants at baseline (Figure 6) ranged from 33.7 to 95.7, with a mean of 50.5, which is lower than total scores for normal development (mean 82.5 to 95.2, depending on gender and age group; Valier 2017). Scores by domain were lowest for the Physical domain, with a mean (SD) score of 32.57 (24.924) and highest for the Emotional domain, with a mean (SD) score of 68.75 (20.283) (data on file). The PedsQL total score (Figure 6) and subdomain scores remained generally stable over time (data on file).

**Figure 6:**
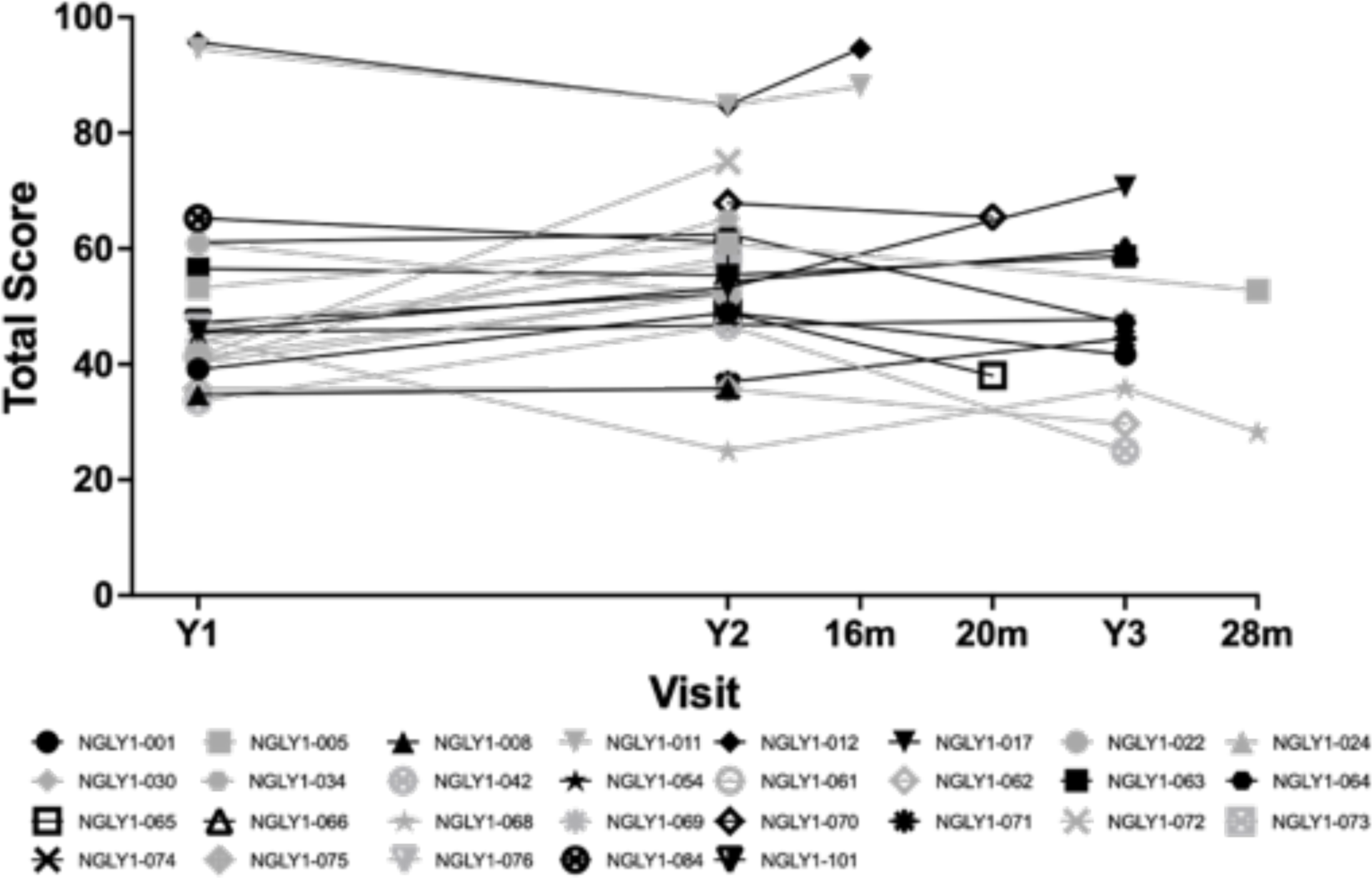
PedsQL (Total Score) vs. Visit. Legend: Black symbols are female and gray symbols are male subjects.

#### Most Bothersome Symptom

When asked to describe the most bothersome symptom of NGLY1 Deficiency, caregivers identified trouble with language/communication (for 13 patients), followed by gross or fine motor deficits, including limited functional hand use (for caregivers of 11 patients) (data on file).

#### GNA levels

GNA levels were elevated in the plasma of all participants, did not correlate with age, and showed minimal change over time (Figure 7). There was no significant linear relationship between age and GNA levels, as indicated by the horizontal black line, suggesting GNA levels (GNA concentration in plasma) remain consistent over time. For onsite participants, mean (SD) GNA plasma concentration was 115 (28.4) ng/mL with range of 57.8 – 189.0 ng/mL, which was 4.3-fold over related controls (unaffected siblings and parents) and 8.5-fold over unrelated unaffected controls (Mueller 2022). Longitudinal GNA samples (n=10) showed ≤ 37 ng/mL change over 2-3 years (max – min concentration; mean change 23.0 ng/mL +/− 10.8 ng/mL).

**Figure 7:**
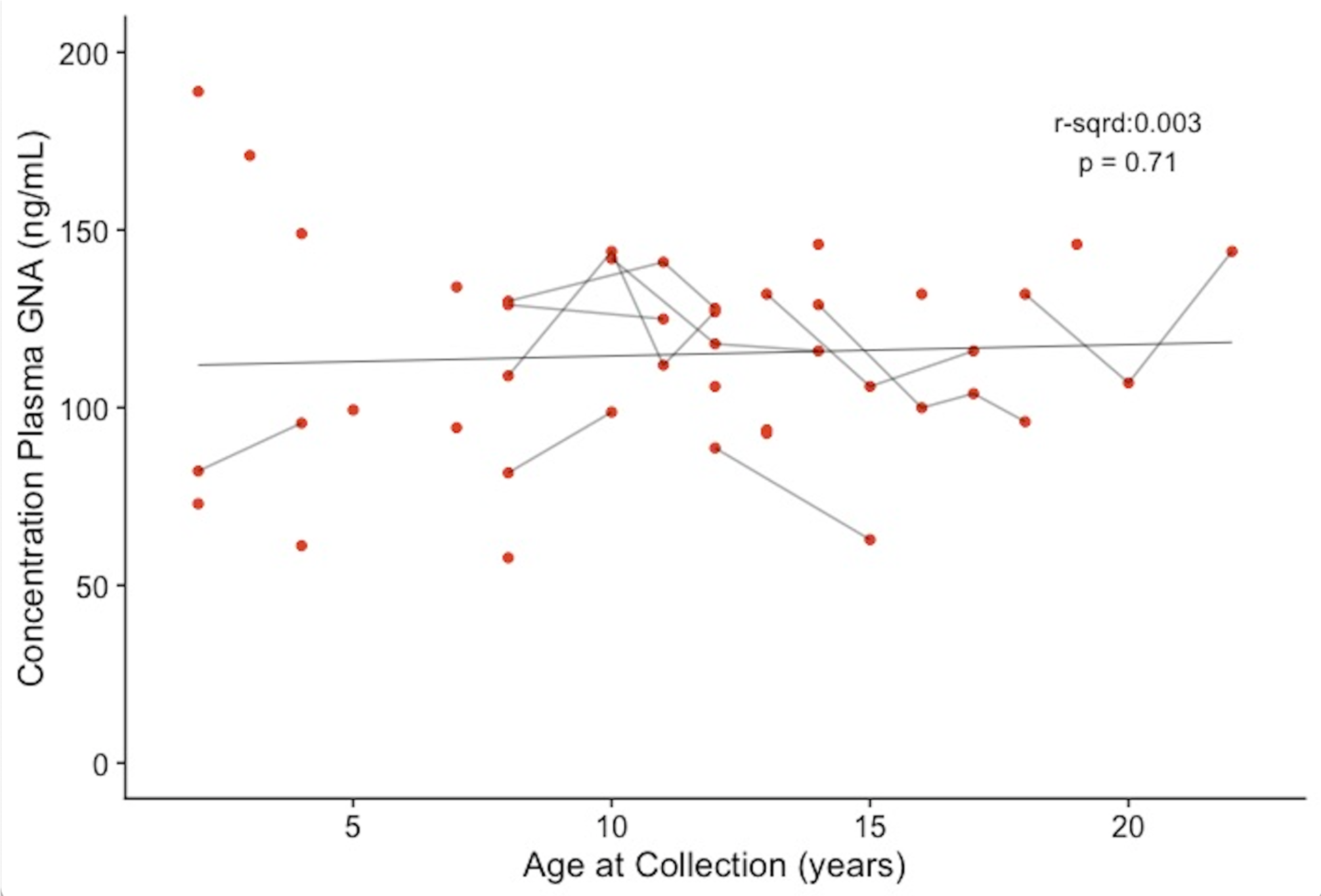
Plasma GNA Concentration vs. Age at Collection for Participants as Measured over Time. Legend: Age at sample collection and GNA concentration were compared to determine whether there was a significant relationship. The line is the trend line with statistical correlation shown. Each red point represents a measurement, and red points with short black lines between them are measurements for the same participant.

#### Laboratory Studies

Laboratory study results were generally unremarkable, except for ALT (Figure 8) and AST (Figure 9) levels, which were elevated in some participants. Those with elevated ALT/AST values showed improvement over time, especially among the younger patients. The liver transaminases were not associated with elevated total bilirubin levels and did not reach levels indicative of severe liver disease.

**Figure 8:**
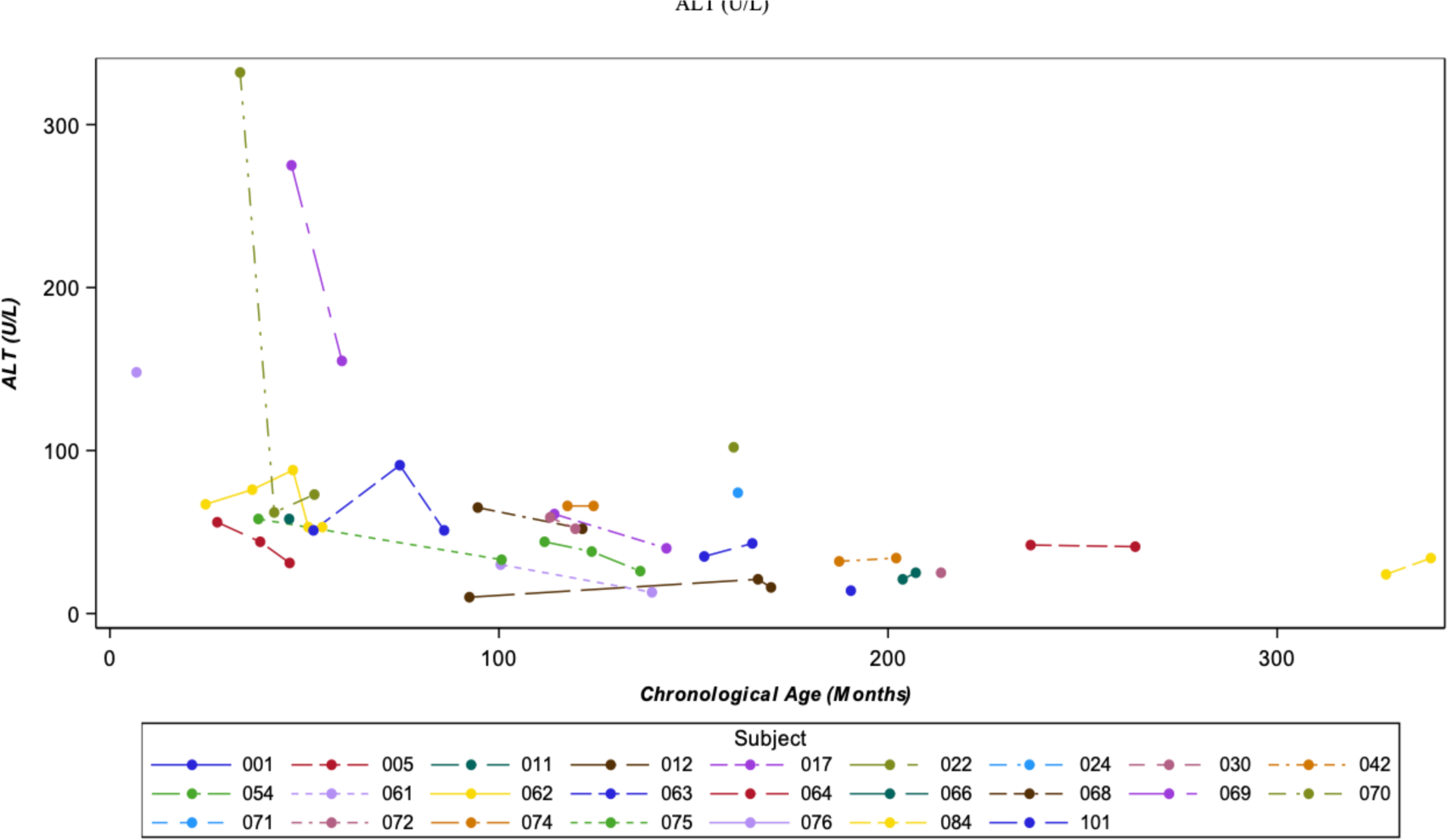
ALT (U/L) vs Chronological Age. Legend: If participants had multiple lab measurements at the same visit, the highest value was selected for analysis.

**Figure 9:**
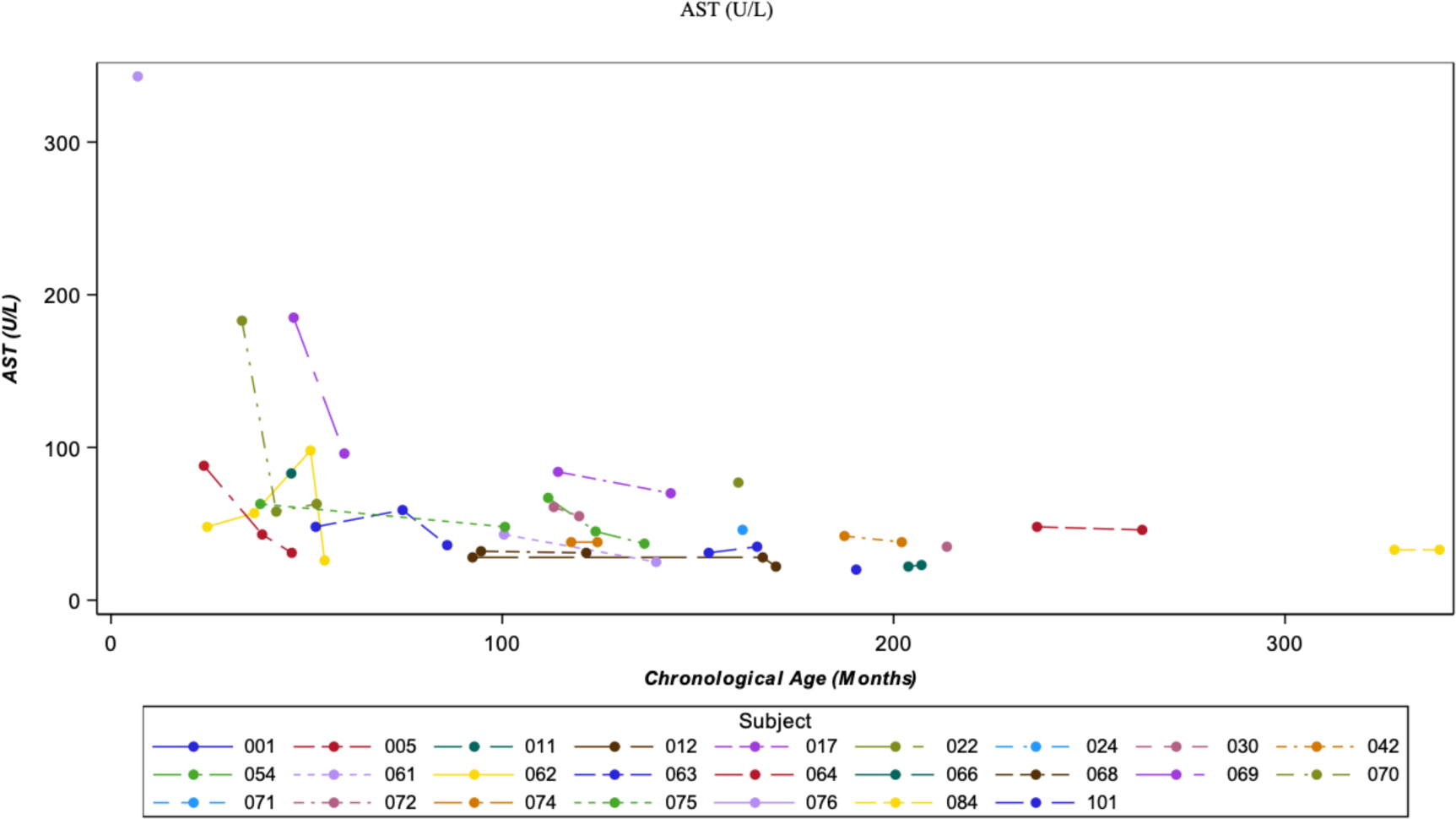
AST (U/L) vs Chronological Age. Legend: If participants had multiple lab measurements at the same visit, the highest value was selected for analysis.

## Discussion

This prospective NHS was undertaken to characterize the clinical characteristics, disease course, and potential clinical trial endpoints for NGLY1 Deficiency, a severely debilitating, ultra-rare, autosomal recessive, neurodevelopmental disorder. Consistent with previous publications, the core features of the disease, global developmental delay and/or intellectual disability, a hyperkinetic movement disorder, transient elevation of transaminases, and (hypo)alacrima (Adams 2018; Enns 2014; Lam 2018; Levy 2022) were found to be profound and did not show spontaneous improvements, as with typical childhood development over time. The natural history was generally slowly progressive or at best stable over at least a 1-year period. This publication reports on neurodevelopmental outcomes, disease course over time, laboratory studies, HRQoL and biomarkers. Mortality is also reported, pointing to the urgency for development of treatments for this devastating disease. This report complements findings on seizures and EEG results previously reported from this NHS (Levy 2022).

All participants in this NHS had profound neurodevelopmental impairment as demonstrated by developmental and adaptive skills assessments using standard validated instruments, the MSEL and Vineland 3. Profound impairment was observed in the composite score and in all domains, encompassing expressive and receptive language as well as coping skills and interpersonal relationships. There was a trend in neurodevelopmental outcomes, such that those participants with epilepsy (>50% of participants) had more severe developmental delay (Levy 2022). While there were minor fluctuations over time with the neurocognitive instruments (MSEL and Vineland 3) across all domains assessed, these were not clinically significant and were generally within the expected variability of measurement. There was no evidence of spontaneous improvements and the participants failed to recover functional abilities typical for their chronological age. Rather, individuals with NGLY1 Deficiency either slowly deteriorated further or remained at a level of profound impairment. In clinical trials of potential therapies for NGLY1 Deficiency, improvements in neurocognitive assessments could signify treatment efficacy; however, even a stabilization or halting of decline based on these assessments could be considered a beneficial effect of treatment.

Nearly all participants exhibited failure to thrive, manifesting as low weight for age, and motor impairments consisting of delayed/absent attainment of motor milestones as shown on physical exam, PDMS-2, and motor subdomains of the MSEL and Vineland-3. Both gross and fine motor skills were severely impaired. Worsening of already poor gross motor function over time was indicated by the reduction in the proportion of participants who achieved unaided sitting or standing with concurrent increased proportions of participants who could not sit or stand. There were no spontaneous improvements in the ability to sit or stand even though participants were at or older than the ages where these gross motor milestones are expected to be achieved. The ability to sit and stand independently would be expected to greatly impact a patient’s and their caregiver’s quality of life. Overall, multiple assessment methods pointed toward slowly progressive declines in motor abilities required for self-care and activities of daily living. The motor function results of this NHS suggest that if motor abilities are used as an endpoint in future therapeutic trials, any improvements seen are more likely to be due to the efficacy of the therapy rather than natural improvement with age, and that even a small improvement would be clinically important, given the profound impairment that begins early in life and may worsen over time.

Schirmer’s test for lacrimation (tear formation) and QSART assessments (sweat response) also indicated impaired functions in participants. These assessments are indicative of autonomic dysfunction in NGLY1 Deficiency (Lam 2018; Pinto 2020; Willems 2022) and were generally stable or worsened over time. Insufficient tear production has an important impact on health in individuals with NGLY1 Deficiency and is associated with corneal damage, including neovascularization, pannus formation, and scarring (Lam 2018). Reduced sweat response (hypohydrosis) requires protective measures against situations that could cause dangerous core body hyperthermia (Lam 2017), such as cooling vests and adequate access to water and air conditioning (Lam 2018).

The majority of participants had impaired HRQoL as measured by PedsQL that showed no clear change over time by total score or by subdomain. Symptoms that were most troublesome to caregivers included difficulties with language/communication and motor deficits (particularly limited functional hand use). The caregivers’ impressions are consistent with the deficits seen on the MSEL, which showed profound impairment in language and communication, the Vineland-3, which showed extensive impairment in gross and fine motor skills, and the PDMS-2, which showed clinically important impairment in all measures of gross and fine motor skills including grasp and object manipulation. Caregiver-reported improvement in the symptoms that most directly affect the HRQoL of participants and their families could represent meaningful secondary endpoints.

GNA has been identified as a substrate biomarker for NGLY1 Deficiency (Mueller 2022). Unlike other potential biomarkers for NGLY1 Deficiency (Lam 2017; Hall 2018; Chang 2019), GNA is a substrate biomarker that directly results from the lack of NGLY1 function. consistently differentiates those individuals affected by NGLY1 Deficiency from those without NGLY1 Deficiency (Mueller 2022). Individuals with the inherited metabolic disease aspartylglucosaminuria (AGU) also show elevated GNA levels but can be distinguished from those with NGLY1 Deficiency through genetic testing and consideration of key clinical features. The data presented in this report demonstrate that GNA was elevated in all NHS participants and remained stable over time. In a rat model of NGLY1 Deficiency, virally-mediated delivery of a functional human copy of the NGLY1 gene led to a reduction in GNA levels that correlated with improvement of behavior and locomotor phenotypes (Mueller 2022, Zhu 2022). A similar restoration of NGLY1 function as assessed by normalization of GNA levels might be used as a key endpoint to measure treatment effect and be expected to predict clinical benefit in future NGLY1 gene therapy clinical trials.

Liver transaminases were mildly elevated at baseline and stable or, in the case of some younger patients, improved over time. This is consistent with previous reports of NGLY1 Deficiency in which liver dysfunction has been identified as an important phenotype of the disease (Need 2012; Heeley 2015; Lipinski 2020; Rios-Flores 2020; Lipari 2020). Although AST/ALT levels were not high enough to be indicative of severe liver disease in this NHS, they should be followed over time during future interventional studies as part of safety monitoring and, concurrently, may be an important clinical measure to evaluate for treatment efficacy (Pandey 2022).

The current study has some limitations. Because patients entered the study at different times, they had different durations of follow-up data. Year 3 study data were only available for a small subset of patients, thus limiting statistical analyses and conclusions about long-term disease progression. The COVID-19 pandemic delayed or precluded some on-site study visits, which impacted the availability of some data. In addition, patients were not able to perform all of the planned assessments, such as the Beery-Buktenica Test of Visual Motor Integration and the 10-MWT, due to the profound neurological and physical impairments associated with NGLY1 Deficiency.

The data presented here provide valuable insights about the clinical course of NGLY1 Deficiency. The sample size of 29 participants in this NHS represents a substantial proportion of the population of approximately 100 identified individuals with NGLY1 Deficiency (Enns 2014; Pandey 2022; data on file). This study demonstrates that the profound neurodevelopmental impairments, quality of life, and other outcomes of NGLY1 Deficiency do not improve and often decline over at least a one-year period, despite existing interventions like physical therapy and symptom management. This dataset informs endpoints, selection of assessment tools, and monitoring intervals for interventional clinical trials. Such assessments include GNA levels as a pharmacodynamic biomarker, instruments that measure language/communication and motor deficits (particularly improved hand use), HRQoL, clinical/caregiver global impressions of change, tear production, and QSART. Seizures and neurophysiologic characterization are also relevant because, as previously described, almost all participants had EEG abnormalities, indicating increased risk of epilepsy (Levy 2022). As the overall disease course shows a slow but progressive decline in neurocognitive and motor function, interventions that result in stabilization or a slowing of the decline in function could be considered as efficacious.

## Data Availability

The datasets that support this study are available from the corresponding author upon reasonable request, subject to institution ethical considerations for participant privacy and consent.

## Acknowledgements

Funding for this study was provided by a grant from Grace Science Foundation (GSF) to Stanford University. Dr Maura Ruzhnikov contributed to protocol development and was the principal investigator of the study. Dr Stephen Maricich reviewed and analyzed preliminary data. Medical writing, editing, and design assistance was provided by Judy Wiles, Trish Rawn, and Selma Tse of Facet Communications Inc. Caroline Stanclift served as a liaison between GSF and the trial site; graphic presentations were provided by Alicia Newton.

## Author contributions

Conceptualization: S.S.D., W.F.M., B.J.B., M.W., K.J.L., J.K.; Study coordination: M.W., C.H.F; Data curation: S.S.D., W.F.M.; Formal analysis: S.T., P.V., S.S.D., W.F.M., C.H.F., S.M; Investigation: S.D.D., W.R.M., J.K., J.P., C.H.F, S.M.; Visualization: S.T., W.F.M., K.J.L.; Writing-original draft: S.T., P.V., S.S.D., W.F.M; Writing-review & editing: S.T., P.V., S.S.D., W.F.M., B.J.B., M.W., K.J.L, J.K., J.P., C.H.F.

## Ethical Approval

Informed consent was obtained from all study participants according to the study protocol approved by the Stanford Institutional Review Board (IRB Protocol # 47335). Parents gave written informed consent for their children or dependents.

